# Inhaled corticosteroids downregulate SARS-CoV-2-related gene expression in COPD: results from a RCT

**DOI:** 10.1101/2020.08.19.20178368

**Authors:** Stephen Milne, Xuan Li, Chen Xi Yang, Ana I Hernandez Cordero, Fernando Sergio Leitao Filho, Cheng Wei Tony Yang, Tawimas Shaipanich, Stephan F van Eeden, Janice M Leung, Stephen Lam, Don D Sin

**Author notes:** authors made equal contributions to the work (Stephen Milne, Xuan Li). **<u>CORRESPONDING AUTHOR</u>:** Dr Stephen Milne UBC Centre for Heart Lung Innovation Rm 166 – St Paul’s Hospital 1081 Burrard Street Vancouver V6Z 1Y6 British Columbia, Canada E.

## Abstract

**Rationale:** Chronic obstructive pulmonary disease (COPD) is a risk factor for severe COVID-19. Inhaled corticosteroids (ICS) are commonly prescribed for the prevention of acute exacerbations in people with COPD, but their use is associated with increased risk of respiratory infections. The effects of ICS on SARS-CoV-2 susceptibility or COVID-19 severity are currently unknown.

**Objectives:** To determine the effects of ICS treatment on the bronchial epithelial cell expression of key SARS-CoV-2-related genes in volunteers with COPD.

**Methods:** We performed a randomized, open-label, parallel treatment trial of 12 weeks treatment with ICS in combination with long-acting beta-agonist (formoterol/budesonide 12/400 µg twice daily or salmeterol/fluticasone propionate 25/250 µg twice daily), or treatment with LABA only (formoterol 12 µg twice daily), in volunteers with mild to very severe COPD. We obtained bronchial epithelial cell samples via bronchoscopy before and after treatment, and determined transcriptome-wide gene expression by RNA sequencing.

**Main Results:** 63 volunteers were randomized to receive treatment. Compared to formoterol alone, formoterol/budesonide treatment decreased the expression of the SARS-CoV-2 receptor gene *ACE2* and the host cell protease gene *ADAM17*. These genes were highly co-expressed with innate immune response genes, particularly those of the type I interferon and anti-viral response pathways, which also tended to decrease following ICS treatment.

**Conclusions:** This is the first randomized controlled trial to show that ICS affect the expression of key SARS-CoV-2-related genes in COPD. Their relation to important anti-viral response genes may have critical implications for SARS-CoV-2 susceptibility or COVID-19 severity in this vulnerable population.

## Introduction

COVID-19, which is caused by infection with the SARS-CoV-2 virus, exhibits a broad spectrum of severity ranging from asymptomatic to severe pneumonia and death. Observational studies show that chronic obstructive pulmonary disease (COPD) is associated with increased risk of severe COVID-19 and mortality (1), the underlying biological mechanisms of which are currently unclear.

Inhaled corticosteroids (ICS) are commonly prescribed as maintenance therapy in COPD. Despite their clinical efficacy in a subset of patients, ICS have been associated with increased risk of bacterial pneumonia in COPD (2) and impaired immune response to viruses *in vitro* (3). There is currently no epidemiological evidence that ICS treatment modifies COVID-19 susceptibility or the severity of infection in COPD patients (4, 5). However, there is poor understanding of how ICS medications affect cellular processes relevant to SARS-CoV-2 biology.

We previously demonstrated that COPD is associated with increased expression of angiotensin-converting enzyme 2 *(ACE2)*, the gene encoding the SARS-CoV-2 receptor, in human lungs (6, 7). Recent *in vitro* studies suggest that ICS may downregulate ACE2 at the gene and protein level (8). This is supported by observational data in both COPD and asthma patients, where ICS use was associated with decreased *ACE2* mRNA in sputum (8, 9). However, the effects of ICS on genes related to SARS-CoV-2 infection and the subsequent host response have not been examined in prospective interventional studies.

In this study, we measured gene expression in lower airway bronchial epithelial cells (BEC) to determine if ICS treatment alters the expression of SARS-CoV-2-related genes (including cell surface receptors, proteases, and innate immune response genes) as part of a randomized controlled trial (RCT) in COPD. We collected the BEC specimens before and after 12 weeks of treatment with long-acting beta-2 adrenoceptor agonist (LABA)/ICS combination therapy or LABA monotherapy. ICS treatment led to relative downregulation of the SARS-CoV-2 receptor gene *ACE2* and the host cell protease gene *ADAM17*. These genes were highly connected to antiviral, innate immune response genes, which may have implications for COVID-19 severity in this vulnerable population.

## Results

### Characteristics of participants in the DISARM trial

In the DISARM study (http://clinicaltrials.gov identifier NCT02833480), we randomized 63 subjects with mild to very severe COPD to receive LABA monotherapy with formoterol (FOR), or LABA/ICS combination therapy with formoterol/budesonide (FOR/BUD) or salmeterol/fluticasone (SAL/FLU), for 12 weeks (Figure 1). Bronchial brush samples were collected from 6–8^th^ generation airways via bronchoscopy before and at the end of treatment. The majority of participants were male and had moderate to severe airflow obstruction (Table 1).

**Figure 1:**
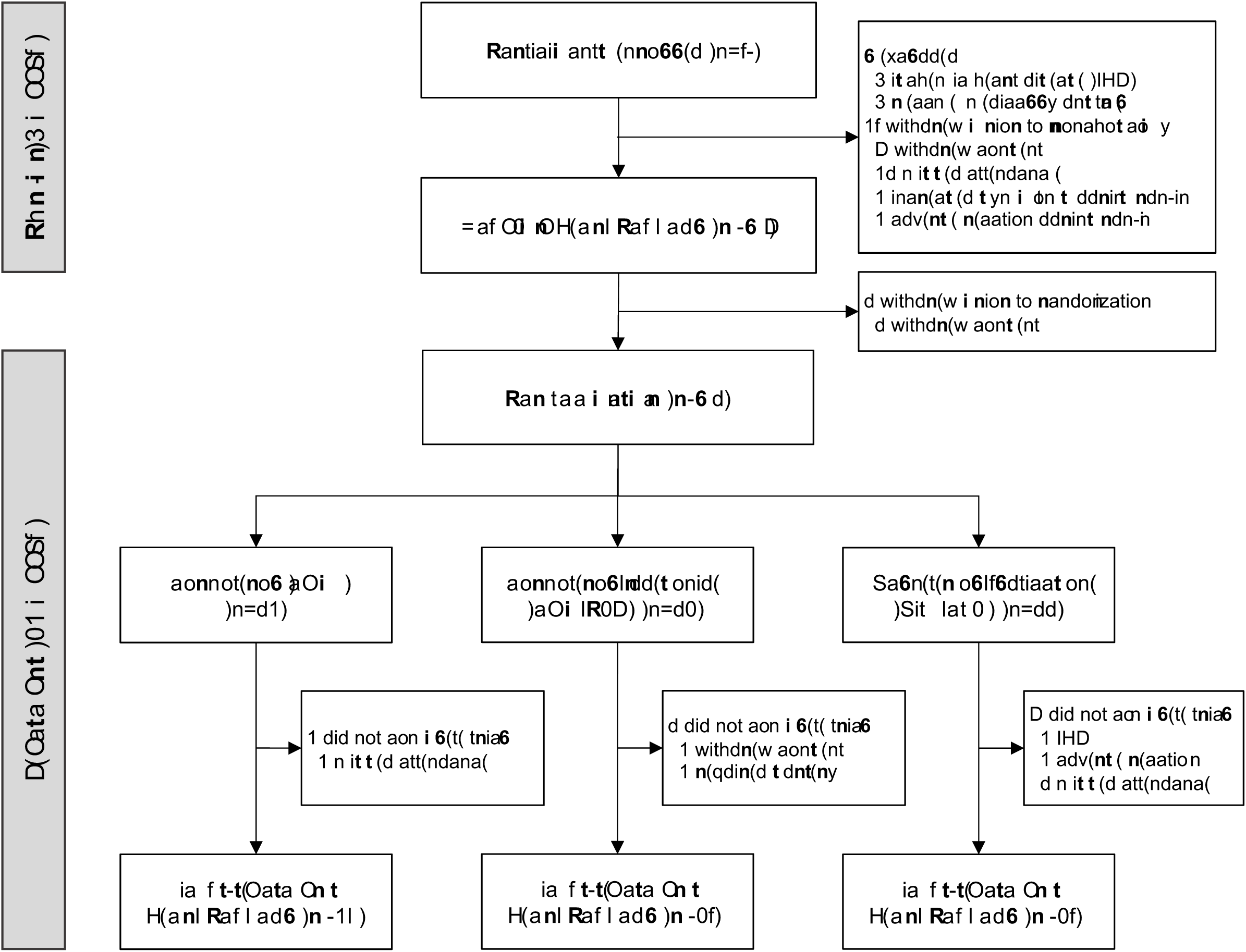
CONSORT flow chart for the DISARM trial. Run-in period comprised 4 weeks treatment with formoterol (FOR) 12 µg inhaled twice daily via Turbuhaler. Randomization was in 1:1:1 ratio in blocks of 6, stratified by prior history of inhaled corticosteroid use. Treatment was open-label with FOR 12 µg inhaled twice daily via Turbuhaler, formoterol/budesonide (FOR/BUD) combination 12/400 µg inhaled twice daily via Turbuhaler, or salmeterol/fluticasone propionate (SAL/FLU) 50/250 µg inhaled twice daily via Diskus, for 12 weeks.

**Table 1:** Characteristics of randomized participants in the DISARM trial. Data are mean (SD) unless otherwise indicated. ^†^obesity defined as BMI≥30 kg/m^2^. Abbreviations: FOR, formoterol; BUD, budesonide; SAL, salmeterol; FLU, fluticasone propionate; IQR, interquartile range; BMI, body mass index; FEV_1_, forced expiratory volume in 1 second; FVC, forced vital capacity; LAMA, long-acting muscarinic antagonist; SGRQ, St George’s Respiratory Questionnaire.

|  | Treatment arm |  |  |
| --- | --- | --- | --- |
|  | FOR | FOR/BUD | SAL/FLU |
| Participants, n | 21 | 20 | 22 |
| Male sex, n (%) | 18 (86) | 17 (85) | 17 (77) |
| Age, median (IQR) years | 63<br>(55 - 68) | 66<br>(63 - 70) | 66<br>(60 - 72) |
| BMI, median (IQR) kg/m<sup>2</sup> | 24.30<br>(21.19 - 30.56) | 23.87<br>(20.28 - 27.22) | 27.25<br>(23.01 - 30.43) |
| Current smoker, n (%) | 11 (52) | 7 (35) | 11 (50) |
| Hypertension, n (%) | 5 (24) | 6 (30) | 4 (21) |
| Obesity<sup>†</sup>, n (%) | 6 (29) | 4 (20) | 6 (27) |
| FEV<sub>1</sub>, median (IQR)<br>%predicted | 64.5<br>(50 - 73) | 58.0<br>(48.1 - 74.5) | 58.0<br>(45.8 - 73) |
| FEV<sub>1</sub>/FVC, median (IQR)<br>% | 56.1<br>(45.3 - 62.8) | 49.0<br>(44.6 - 61.5) | 52.3<br>(48.8 - 64.4) |
| GOLD stage I/II, n (%) | 13 (62) | 11 (55) | 12 (55) |
| GOLD stage III/IV, n (%) | 8 (38) | 9 (45) | 10 (45) |
| Respiratory medications |  |  |  |
| LAMA, n (%) | 19 (91) | 19 (95) | 16 (73) |
| Methylxanthines, n (%) | 0 (0) | 1 (5) | 0 (0) |
| Health status (SGRQ) |  |  |  |
| Symptom domain | 44.5 (23.5) | 47.5 (19.8) | 49.9 (26.3) |
| Activity domain | 61.1 (13) | 60.3 (27.8) | 58.9 (19.1) |
| Impact domain | 29.5 (15.8) | 26.6 (15.9) | 26.0 (20.2) |
| Total score | 41.7 (14.8) | 39.4 (17.6) | 39.7 (19.2) |
| Peripheral blood cells |  |  |  |
| White blood cells<br>(x10<sup>9</sup>/mL) | 7.92 (2.24) | 6.80 (2.35) | 7.73 (1.77) |
| Neutrophils (x10<sup>9</sup>/mL) | 5.12 (1.86) | 4.46 (1.83) | 4.41 (1.12) |
| Eosinophils (x10<sup>9</sup>/mL) | 0.24 (0.12) | 0.24 (0.15) | 0.27 (0.11) |

### ICS treatment downregulates key SARS-CoV-2 related genes in COPD

In order to quantify BEC gene expression, we performed whole-transcriptome RNA sequencing (RNA-seq) on bronchial brush samples. After quality control filtering, a total of 15,263 genes were included in the expression dataset. We examined the expression levels of key SARS-CoV-2-related genes: *ACE2* (encodes the putative SARS-CoV-2 entry receptor ACE2) (10); *BSG* (encodes an alternative entry receptor basigin, also known as CD147) (11); *TMPRSS2* (encodes a cell surface metalloproteinase that primes the SARS-CoV-2 spike protein to facilitate viral entry) (10); *ADAM17* (encodes a metalloproteinase that cleaves the ACE2 protein and facilitates endocytosis of the ACE2-SARS-CoV-2 complex) (12); and *FURIN* (encodes the subtilisin-like peptidase furin, which can prime the SARS-CoV-2 spike protein) (13, 14).

Sixty one participants had pre-treatment gene expression data available. There were no obvious differences in overall gene expression between the treatment groups at baseline (Figure E1). Each of the key SARS-CoV-2-related genes were expressed in pre-treatment BECs (Figure 2A). Pre-treatment *ACE2* and *BSG* expression were negatively associated with age (p = 3.4×10^−4^ and p = 0.033, respectively; Figure 2B). Compared to ex-smokers, current smokers had significantly greater pre-treatment expression of *ACE2* (p = 9.3×10^−5^), *BSG* (p = 4.8×10^−4^) and *TMPRSS2* (p = 0.028) (Figure 2C).

**Figure 2:**
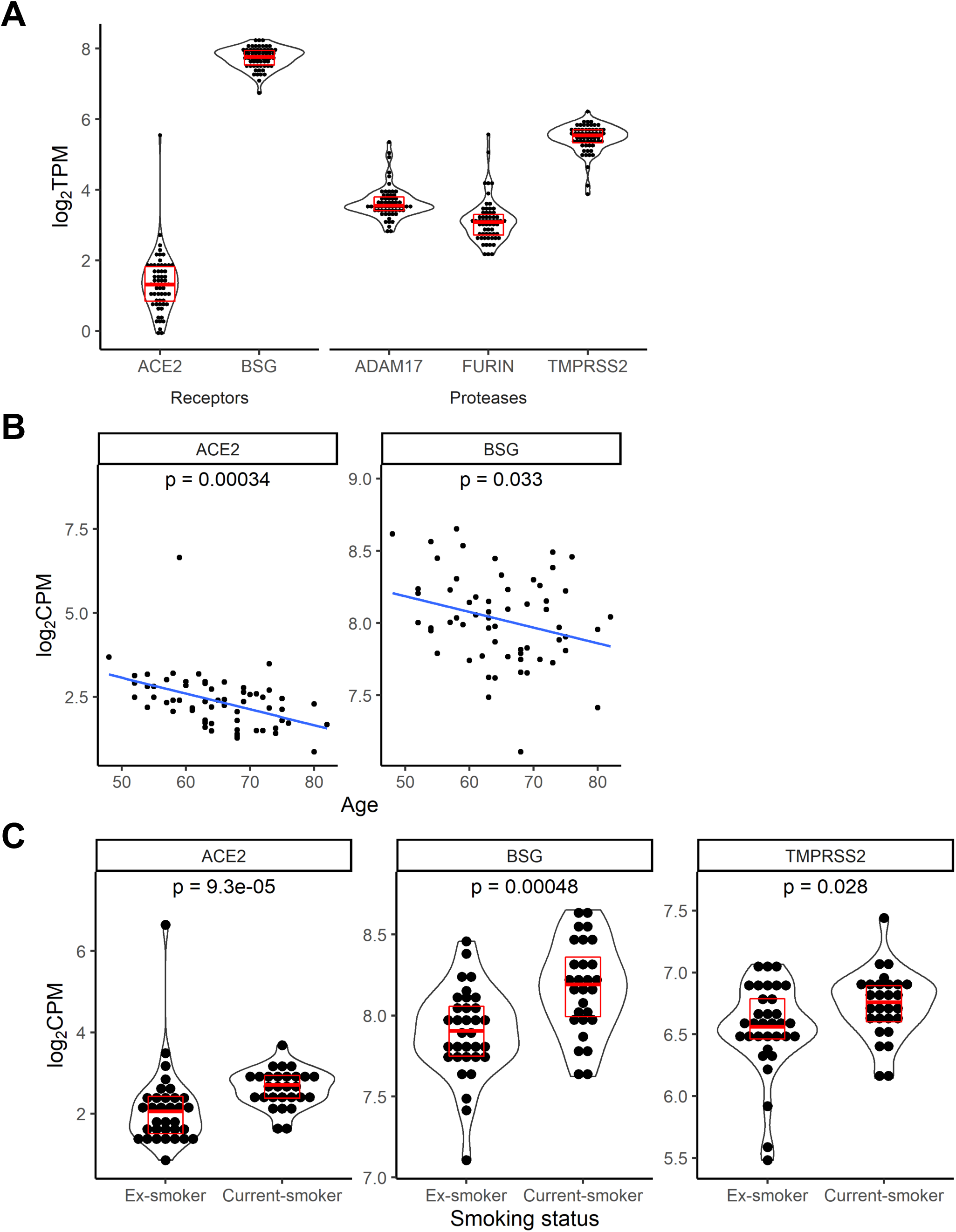
Baseline (pre-treatment) bronchial epithelial cell expression of SARSCoV-2-related genes in COPD. Total samples n = 61 (two out of the 63 randomized participants had insufficient RNA for sequencing). (A) Each of the 5 key SARS-CoV-2-related genes were expressed at baseline. (B) Increasing age was associated with decreasing expression of *ACE2* and *BSG*. P value from linear regression (C) Current smokers had significantly greater expression of *ACE2, BSG* and *TMPRSS2* genes compared to ex-smokers. p value from Wilcoxon rank-sum test. Abbreviations: TPM, transcripts per million; CPM, counts per million.

Fifty four participants had both pre- and post-treatment gene expression data available. Compared to baseline levels, LABA-only treatment with FOR significantly increased expression of *ACE2* (median Δlog_2_CPM +0.47, p = 0.002) and *ADAM17* (median Δlog_2_CPM +0.13, p = 0.03). LABA/ICS combination treatment with FOR/BUD significantly decreased *ADAM17* expression (median Δlog_2_CPM –0.19, p = 0.003), while treatment with SAL/FLU did not significantly alter expression of any of the key genes (Table E1, Figure E2).

A placebo arm was not included in the DISARM trial as this could predispose participants to a worsening of their COPD symptoms or acute exacerbation. However, we were able to isolate the effects of ICS therapy by comparing the pre- to post-treatment changes in each of the LABA/ICS groups to the LABA-only group. Δlog_2_CPM *ACE2* for FOR/BUD and SAL/FLU groups were significantly lower than that of the FOR group (p = 0.049 and p = 0.03, respectively), while Δlog_2_CPM *ADAM17* for FOR/BUD was significantly lower than that of the FOR group (p = 7.05×10^−5^) (Figure 3). These results indicate that ICS have a suppressive effect on the transcription of these genes.

**Figure 3:**
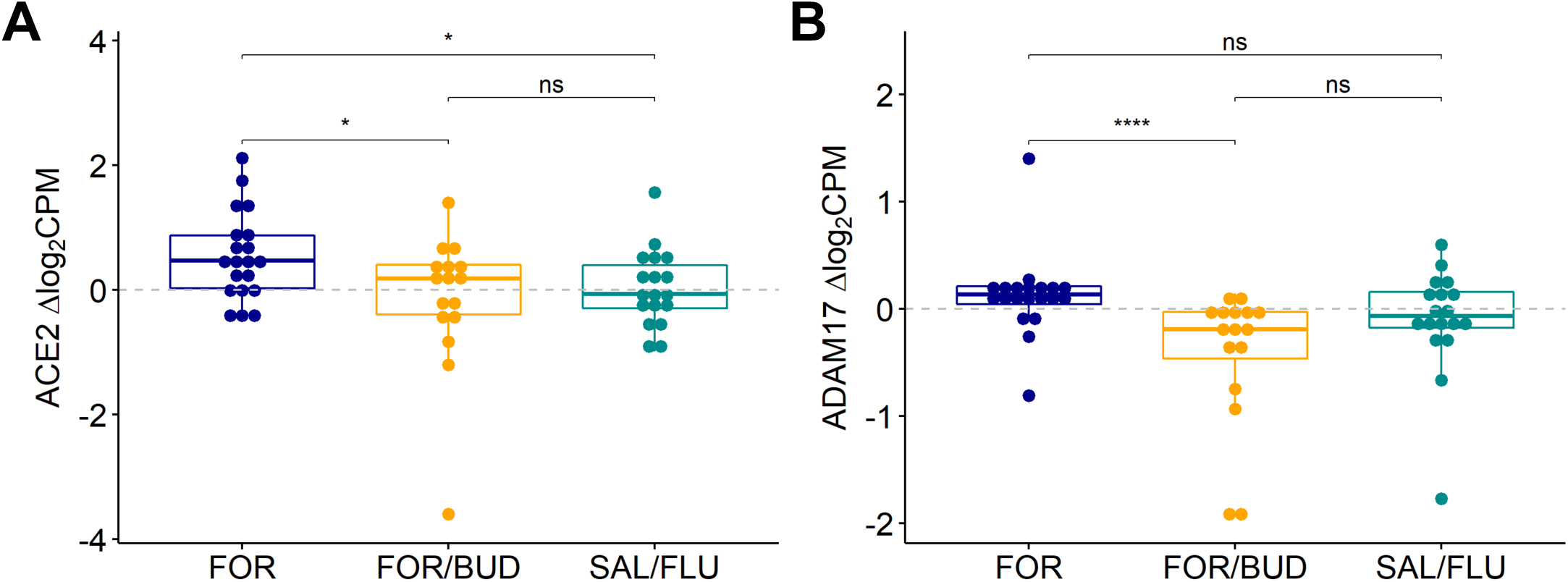
Pre- to post-treatment change in bronchial epithelial cell gene expression (Δlog_2_CPM). Total n = 54 participants had both pre- and post-treatment RNA sequencing data available. Compared to treatment with FOR alone, FOR/BUD and SAL/FLU treatment significantly decreased expression of the SARS-CoV-2 receptor gene *ACE2* (A), and FOR/BUD treatment significantly decreased expression of the host cell protease gene *ADAM17* (B). Between-group comparisons are by Wilcoxon rank-sum test. Abbreviations: FOR, formoterol; BUD, budesonide; SAL, salmeterol; FLU, fluticasone propionate; CPM, counts per million; ns, non-significant. *p< 0.05 ***p< 0.001.

### Transcriptome-wide effects of ICS treatment in COPD

Corticosteroids have pleiotropic effects on gene expression, including positive and negative regulation of transcription (15). We therefore wanted to explore the effects of treatment on the whole transcriptome. After a gene selection process based on pairwise comparisons of Δlog_2_CPM between treatment groups (see Methods Supplement), a total of 1,188 genes were plotted in a clustered heat map (Figure 4). Overall, the changes in gene expression were modest, with the majority of genes having median Δlog_2_CPM within a range of ±1. In the LABA/ICS treatment groups, the direction of effect was evenly split between upregulation (i.e. Δlog_2_CPM > 0) and downregulation (i.e. Δlog_2_CPM < 0). Individual genes tended to show opposite effects between the LABA-only and LABA/ICS treatment arms such that genes upregulated by FOR treatment were downregulated by FOR/BUD and SAL/FLU treatment and vice versa. Changes were mostly consistent between the FOR/BUD and SAL/FLU groups, suggestive of an ICS class effect.

**Figure 4:**
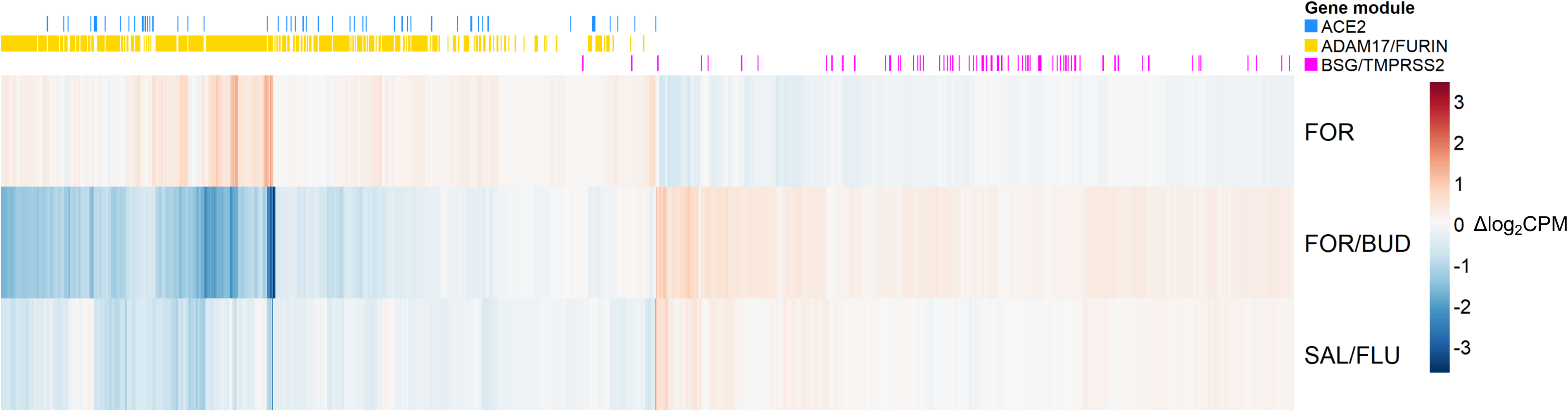
Heat map of pre- to post-treatment change in gene expression (Δlog_2_CPM) across the transcriptome. Each column represents a single gene (total 1,188 genes, selected for plotting based on between-group pairwise Wilcoxon rank-sum test at FDR< 0.05). Treatment with FOR/BUD and SAL/FLU tended to decrease the expression of genes belonging to the *ACE2* and *ADAM17*/*FURIN* coexpression modules. Genes are arranged using hierarchical clustering. Each gene is annotated by membership of one of the SARS-CoV-2-related gene co-expression modules. Abbreviations: FOR, formoterol; BUD, budesonide; SAL, salmeterol; FLU, fluticasone propionate; CPM, counts per million.

### SARS-CoV-2-related genes are co-expressed with innate immune response genes

Co-expression of genes (i.e. correlation between their transcriptional levels) can provide insights into the biological functions of genes that are ‘active’ at the same time (16). We therefore determined genes that were co-expressed with each of the key SARS-CoV-2-related genes in the pre-treatment expression data by performing a weighted gene correlation network analysis (WGCNA) (17, 18). The key SARS-CoV-2-related genes were contained within three of the 20 co-expression modules identified (Figure 5, Tables E2-E4). *ACE2* was in a module of 538 genes; its ‘hub gene’ (i.e. the gene with the strongest correlation to the module eigengene or ‘module membership’) was *DDX58* (DExD/H-box helicase 58), which encodes a protein that is involved in viral dsRNA recognition. *BSG* and *TMPRSS2* were found within a module of 1,528 genes, with the hub gene *AASDHPPT* (aminoadipate-semialdehyde dehydrogenase-phosphopantetheinyl transferase), which encodes a protein involved in amino acid metabolism. *ADAM17* and *FURIN* were found in a module of 2,091 genes with the hub gene *GNA13* (G-protein subunit alpha-13), which encodes a component of G-proteins that are involved in transmembrane signal transduction.

**Figure 5:**
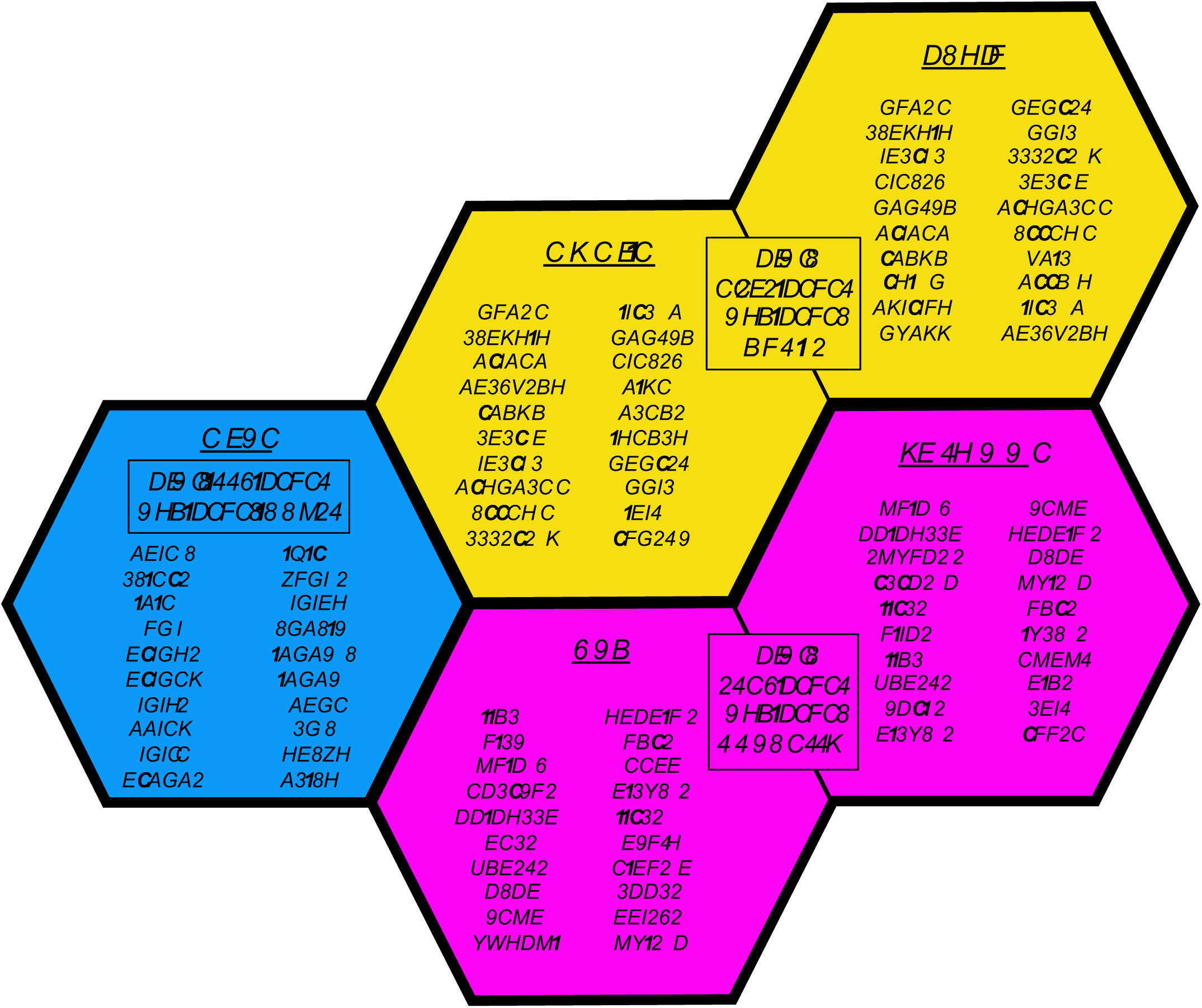
Weighted gene correlation network analysis. This methodology identifies clusters or modules of highly-connected genes. The 5 key SARS-CoV-2-related genes were found in 3 modules, represented by colors. ‘Size’ is number of genes connected to the module eigengene at false discovery rate < 0.05. ‘Hub gene’ has the strongest module membership (i.e. strongest correlation with the module eigengene). Listed are the 20 genes with the strongest connection (based on the weighted topological overlap matrix) to the relevant SARS-CoV-2-related gene. See Zhang and Horvath (17) for full description of the WGCNA methodology and terminology.

To functionally characterize each of the relevant gene modules, we examined the overlap of module membership with Gene Ontology (GO) biological processes (19) at a false discovery rate < 0.05 (Figure 6, Tables E5-E7). The *ACE2* module was highly enriched for genes related to type I interferon (IFN-I) production and response, the response to interferon-gamma, and the response to viruses in general. The *ADAM17*/*FURIN* module was enriched for genes related to the regulation of innate immunity, leukocyte function (including T cell activation and myeloid cell differentiation) and cytokine production. The *BSG*/*TMPRSS2* module was enriched for genes related to cellular processes such as protein transmembrane transport, peroxisome organization, and nucleic acid processing.

**Figure 6:**
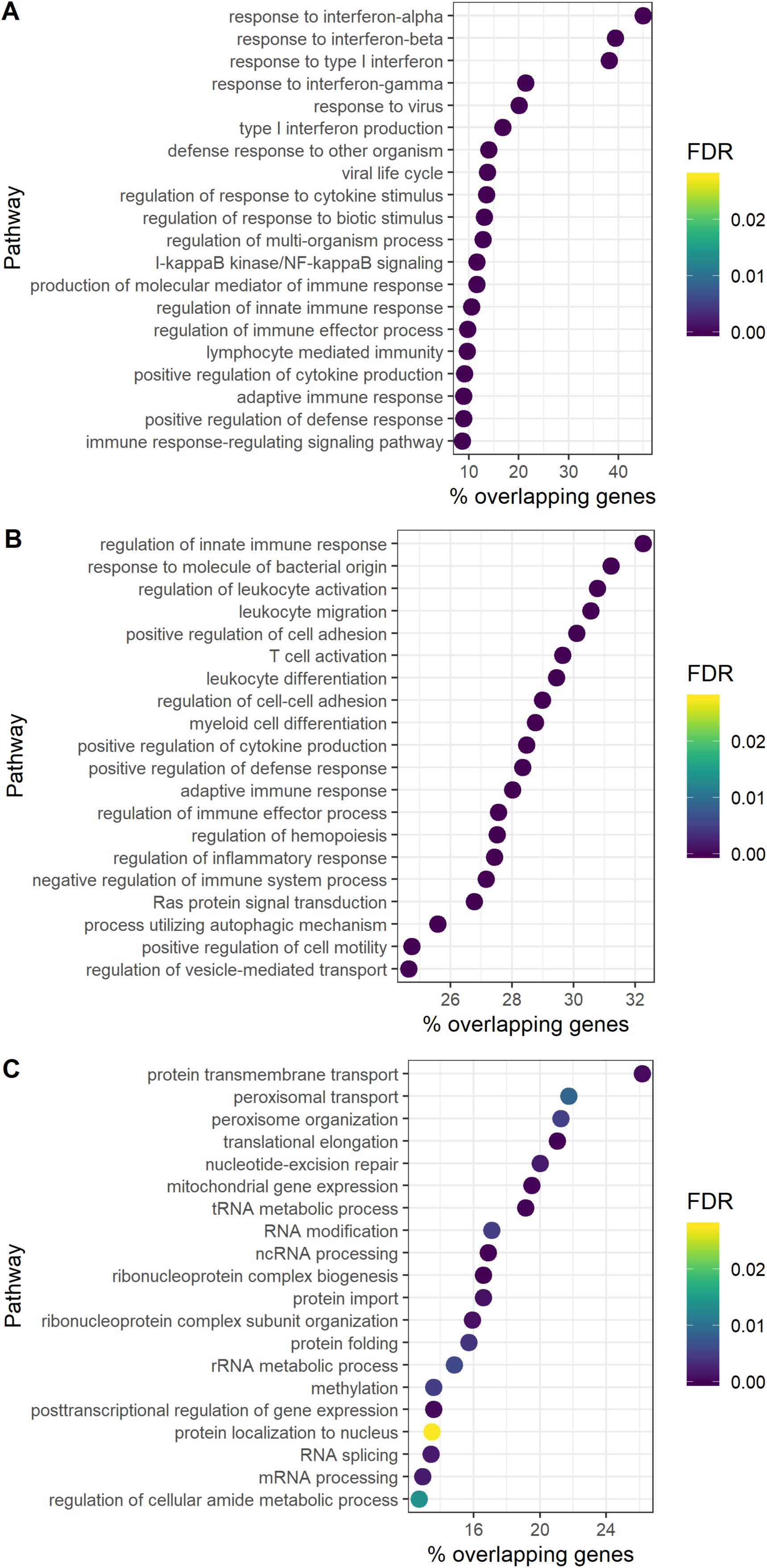
Gene Ontology (GO) biological process enrichment in SARS-CoV-2-related gene co-expression modules. Top 20 GO processes by FDR are shown, ordered by % overlapping genes (proportion of genes in the GO process that also appear in the gene module). The *ACE2* module (A) was highly enriched with genes related to the response to interferons and response to virus. The *ADAM17*/*FURIN* module (B) was enriched with genes related to regulation of innate immunity, leukocyte function, and cytokine production. The *BSG*/*TMPRSS2* module (C) was enriched for genes related to cellular functions including nucleic acid processing. Abbreviations: FDR, false discovery rate.

We next examined the enrichment of the modules with genes in human disease pathways using the Kyoto Encyclopedia of Genes and Genomes (KEGG) (20) (Figure E3, Table E8-E10). The *ACE2* module was enriched for genes in KEGG pathways related to viral infections (including influenza A, measles, Epstein-Barr virus, and herpes simplex virus) and immune-mediated diseases (including primary immunodeficiency, allograft rejection, and type 1 diabetes mellitus). The *ADAM17*/*FURIN* module was enriched for genes related to infectious diseases (including leishmaniasis, *Staphylococcus aureus* infection, and tuberculosis), as well as autoimmune diseases (including rheumatoid arthritis and inflammatory bowel disease). The *BSG*/*TMPRSS2* module was not enriched for any KEGG pathways related to human disease at FDR < 0.05.

Together, these results show that the SARS-CoV-2 receptor gene *ACE2* and the host cell protease genes *ADAM17* and *FURIN* are highly connected to genes of the immune system, particularly those related to the innate immune response to viral infection.

### Genes co-expressed with SARS-CoV-2-related genes are downregulated by ICS treatment

To further explore how ICS treatment affects SARS-CoV-2-related genes, we annotated genes in the heatmap that belonged to each of the three SARS-CoV-2-related gene co-expression modules (Figure 4); 42 percent of the genes in the heat map belonged to one of the SARS-CoV-2-related gene modules. The *ADAM17*/*FURIN* module was over-represented in the heat map (32 percent of mapped genes) compared to the *ACE2* module (4 percent of mapped genes) and *BSG*/*TMPRSS2* module (6 percent of mapped genes). *ADAM17*/*FURIN* and *ACE2* module genes were clustered in areas of the heat map characterized by genes whose expression was downregulated by LABA/ICS.

### Changes in SARS-CoV-2-related gene expression are associated with changes in innate immune response genes

ICS have been shown to alter the expression of innate immune response pathways, including key anti-viral response genes and pro-inflammatory cytokines (3, 15, 21, 22). To further explore how ICS affect these pathways, we examined pre- to post-treatment changes in expression of a panel of genes encompassing virus recognition, IFN-I/III signaling, and inflammatory cytokines/chemokines (Figure 7A). At FDR< 0.05, FOR/BUD treatment significantly reduced the expression of 8 genes in the panel *(IL10RA, IRF3/8, IL1B, IL1R2, CCL3/5* and *CXCL8)*, while SAL/FLU reduced the expression of a single gene *(CCL5)*. We then examined how changes in the key SARS-CoV-2 genes were related to changes in these immune response genes using Pearson correlation (Figure 7B, Figure E4, Table E11). Notably, changes in *ACE2* expression were correlated with changes in the expression of viral sensor genes (e.g. *DDX58, TLR3)*, IFN-I/III signal transducers (e.g. *JAK2, STAT1/2, IRF9)* and interferon-stimulated genes (ISGs) (e.g. *OAS2/3, CXCL10, RSAD2* and *EIF2AK2)* but not inflammatory cytokines/chemokines or their receptor genes. Changes in *ADAM17* expression were correlated with changes in most of the innate immune response genes that were examined, including inflammatory cytokines/chemokines. Together with the WGCNA gene co-expression analysis, these findings show that the expression levels of SARS-CoV-2-related genes are closely related to those of innate immune response genes, suggesting they may share common transcriptional regulation that is modified by ICS.

**Figure 7:**
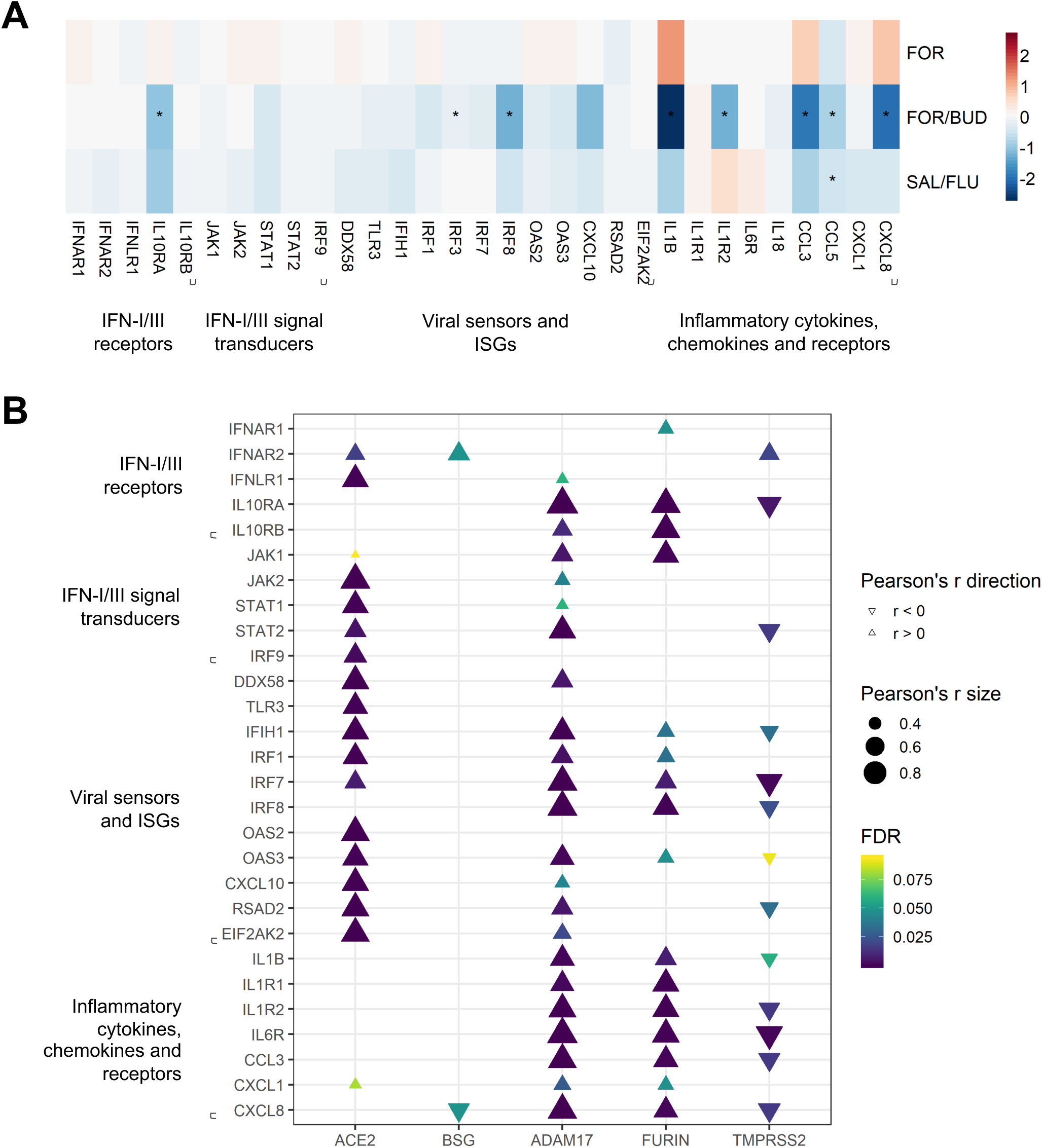
Change in selected innate immune response genes following treatment. (A) Heat map of pre- to post-treatment change in gene expression (Δlog_2_CPM) of innate immune response genes. FOR/BUD and SAL/FLU treatment tended to decrease the expression of the innate immune response genes. *FDR< 0.05 for pre- to post-treatment Wilcoxon signed rank test. (B) Pearson correlation of change in expression (Δlog_2_[TPM+1]) of immune response genes with change in expression of key SARS-CoV-2-related genes following LABA/ICS treatment (FOR/BUD and SAL/FLU groups combined). Changes in *ACE2* expression were highly correlated with changes in IFN-I/II signal transducers, viral sensors and ISGs. Only correlations with FDR < 0.1 are shown. Abbreviations: FOR, formoterol; BUD, budesonide; SAL, salmeterol; FLU, fluticasone propionate; CPM, counts per million; TPM, transcripts per million; IFN-I/III, type I and III interferons; ISGs, interferon stimulated genes

## Discussion

To our knowledge, this is the first prospective RCT in patients with COPD to report the effects of ICS on SARS-CoV-2-related gene expression. We showed that 12 weeks of treatment with combination LABA/ICS (compared to LABA monotherapy) resulted in a significant reduction in the expression of the SARS-CoV-2 receptor gene *ACE2* and the host cell protease gene *ADAM17* in the airway mucosa of COPD patients. Using network analysis, we also showed that *ACE2, ADAM17* and the host cell protease gene *FURIN* were co-expressed with genes involved in the innate immune response and that, qualitatively, these co-expressed genes were also downregulated by ICS therapy. Finally, we showed that changes in the expression of SARS-CoV-2-related genes were correlated with changes in expression of innate immune genes, particularly those of the IFN-I pathway. These results provide an insight into the complex changes induced by ICS in the respiratory epithelium that might impact on the risk of COVID-19 or its outcomes in patients with COPD.

Previous studies suggest that corticosteroid use is associated with an increased risk of infection with common respiratory virus and impaired viral clearance (21). There is also a well-documented association between ICS use and increased risk of pneumonia in COPD (2), which naturally raises concerns that ICS use may be harmful in COPD patients with COVID-19. On the other hand, several findings point towards a potential protective effect of corticosteroids in COVID-19. For example, asthma is more commonly treated with ICS but, unlike COPD, is not associated with increased risk of severe COVID-19 (23). A recent RCT found that systemic dexamethasone treatment improved mortality in severe COVID-19 patients who were mechanically ventilated (24). Additionally, the ICS ciclesonide has been shown to inhibit SARS-CoV-2 replication *in vitro* (25). However, a recent analysis of 148,488 COPD patients in the United Kingdom found no significant association between ICS therapy and COVID-19 severity or mortality (4). Current international consensus statements recommend that patients continue their current COPD medications until further evidence comes to hand (26).

SARS-CoV-2 achieves host cell entry primarily via membrane-bound ACE2. We previously showed that *ACE2* gene and protein expression was increased in the bronchial epithelium (6) and lung tissue (7) of people with COPD, a finding confirmed by other investigators (27, 28). Increased availability of ACE2 may increase the susceptibility to infection and the risk of severe COVID-19 (29, 30). In the present study, *ACE2* expression increased after 12 weeks of formoterol monotherapy, but in the absence of a placebo arm it is unclear if this represents an effect of the intervention or some other driving force. Nevertheless, the addition of ICS in the form of FOR/BUD and SAL/FLU significantly attenuated this increase, suggesting that ICS is active in the regulation of *ACE2* expression. If high expression of *ACE2* increases the susceptibility of cells to SARS-CoV-2 infection, then downregulation by ICS should, in theory, be protective. However, since ACE2 is a critical counter-regulator of the renin-angiotensin-aldosterone system, its downregulation may predispose to angiotensin-II/AT1-mediated acute lung injury (ALI). This phenomenon has been demonstrated in *Ace2* knockout mouse models of aspiration-, sepsis- and SARSCoV-induced ALI (31, 32).

Current evidence suggests that *ACE2* expression is intrinsically linked to the activity of IFN-I. Recently, Ziegler et al (33) showed that ACE2 expression in primary nasal epithelial cells was induced by IFN-alpha, suggesting *ACE2* is an interferon-stimulated gene (ISG) in epithelial cells. Our results support this hypothesis in two ways: first, using WGCNA, we found that *ACE2* co-expressed genes were highly enriched with genes involved in the IFN-I pathway; and second, the changes in *ACE2* induced by ICS were highly correlated with changes in other ISGs such as *OAS2*/3, *CXCL10*, and *RSAD2* (34). The IFN-I pathway is critical in the early cellular response to viral infection, acting as a ‘first alarm’ by inducing the expression of ISGs (which create an anti-viral state within the cell) and pro-inflammatory cytokines (34). The function of *ACE2* as an ISG is unknown, but its induction by IFN-I may serve to counteract any accompanying angiotensin-II/AT1-mediated lung inflammation following viral infection.

Coronaviruses have a particular ability to evade the IFN response, which allows efficient and rapid viral replication (35). However, outcomes for the host may depend on the extent and timing of the IFN response mounted. In a murine model of SARSCoV infection, Channappanavar et al (36) showed that a robust early IFN response limited viral replication and prevented excessive lung damage, while a robust but delayed IFN response was associated with excessive inflammation and death. Interestingly, a muted or absent IFN-I response was associated with minimal lung damage despite high viral titers (36) highlighting the potential pathogenic role of the IFN-I response. Other recent studies support the hypothesis that IFNs contribute to COVID-19 pathology (37, 38). Zhuang et al recently determined that SARS-CoV-2 can positively induce *ACE2* expression, most likely by activating IFN-I pathways (39). We speculate that *ACE2* upregulation by IFNs may result in a feed-forward cycle whereby the anti-viral response of epithelial cells actually enhances their susceptibility to SARS-CoV-2 infection. Thus the IFN response to SARS-CoV-2 may be a two-edged sword: an early and appropriately-regulated IFN response may inhibit the virus and cause minimal symptoms, whereas a delayed or dysregulated IFN response may facilitate viral replication, dissemination, and excessive inflammation leading to severe COVID-19. Consistent with this hypothesis, recent studies in people with COVID-19 support a role for treatment with IFN-I if administered early in the clinical course (40, 41). COPD patients already have impaired IFN-I responses following viral infection (42), meaning this patient population may be particularly susceptible to severe COVID-19 by this mechanism and thus candidates for targeted, early IFN treatment.

ICS may therefore affect COVID-19 outcomes by modifying IFN signaling in the airway epithelium. *In vitro* studies have shown that corticosteroids decrease IFN-I expression (21, 43). However, consistent with previous findings in COPD, the expression levels of these genes in our BEC samples were extremely low and therefore we cannot confirm any treatment effect. Regardless, how ICS treatment affects the magnitude and timing of the IFN-I responses following viral infection may be more important, and this should be the focus of future investigations.

We also showed that LABA/ICS therapy significantly downregulated BEC expression of *ADAM17*. The viral S protein induces ADAM17-dependent shedding of the ACE2 ectodomain, creating the soluble form of ACE2 and facilitating fusion of the viral and cell membranes (44). Inhibition of ADAM17 at least partially blocks SARS-CoV entry in cultured epithelial cells (12, 44), highlighting this protease as a potential therapeutic target. ADAM17 also plays a crucial role in interleukin-6 (IL-6) signaling, which is activated in severe COVID-19 (45). Shedding of the IL-6 receptor (IL-6R) ectodomain facilitates a pro-inflammatory response via the so-called *trans*-signaling IL-6 pathway (46), and selective blockade of IL-6 *trans*-signaling can attenuate sepsis in mice (47). ADAM17 has therefore been described as a “master switch” between the pro-inflammatory *trans*- and anti-inflammatory *classical*- (i.e. via membrane-bound IL-6R) IL-6 signaling pathways (48). However, since inhibitors of ADAM17 have not been successful in clinical trials of inflammatory diseases (49), a beneficial effect of *ADAM17* downregulation by ICS on COVID-19 outcomes is, at this time, speculative.

There are several limitations to our study. First, the relatively small sample size may explain why there were many non-significant trends in pre- to post-treatment change in gene expression. However, the largely consistent directions of change between ICS treatment arms suggests these effects were not due to chance alone. Second, we do not have longer-term follow-up gene expression data to know if the observed changes are durable. Third, we enrolled only stable participants so cannot know if ICS alters the expression of these genes during unstable periods. Fourth, we studied gene rather than protein expression and so cannot necessarily infer changes in protein levels or function. Finally, our data were derived from BECs rather than alveolar or endothelial cells, and may not directly reflect the risk of SARS-CoV-2 infection or ALI in the lung.

Our results show that ICS medications have activity on the expression of key SARSCoV-2 related genes in COPD patients, and that expression of the SARS-CoV-2 receptor gene *ACE2* is closely related to the expression of anti-viral IFN-I pathways. Whether this translates into altered COVID-19 outcomes requires further study: there are at least two RCTs currently underway (http://clinicaltrials.gov identifiers NCT04381364 and NCT04355637) which may shed some light. Ultimately, optimal control of COPD symptoms and exacerbation risk may be the most important determinants of clinical outcomes in these patients. For this reason, we agree with the international consensus that ICS use in COPD patients should be continued if clinically indicated until further evidence is available.

## Methods

Comprehensive methods are presented in the Online Data Supplement.

## Design of the DISARM trial

The DISARM study aimed to determine the effects of ICS on the lung microbiome in COPD. The full trial protocol is registered at http://clinicaltrials.gov (NCT02833480) and the study was approved by the Human Research Ethics Committee of the University of British Columbia and Providence Health Care (H14–02277). After providing written informed consent, enrolled participants with mild to very severe COPD went through an initial 4-week run-in period with FOR plus short-acting beta-agonist (SABA) as required followed by a baseline bronchoscopy. They were then randomized to receive ongoing LABA monotherapy with FOR, or LABA/ICS combination therapy with FOR/BUD or SAL/FLU for 12 weeks, followed by a second bronchoscopy. At each bronchoscopy, bronchial brushes (6–8^th^ generation airway) and bronchoalveolar lavage (right middle lobe or lingula) were collected from sites away from any pathology identified on CT scan. Bronchoscopists and laboratory personnel were blinded to treatment assignment.

## Sample processing, RNA extraction and RNA sequencing

RNA was extracted from cytological brush specimens stored in QIAzol RNA lysis buffer (QIAGEN, Stockach, Germany) using the RNeasy Plus kit (QIAGEN) and sample quality control was performed using the Agilent 2100 Bioanalyzer (Agilent, Santa Clara, CA, USA). RNA sequencing was performed on the Illumina NextSeq 500 (Illumina, San Diego, CA, USA) with paired end 42bp-by-42bp reads. RNA-seq data is available at [add prior to publication].

## Bioinformatics and statistical comparisons

Quality control of raw sequencing reads, alignment, and count normalization procedures are described in the Online Data Supplement. Principal component analysis was used to visualize data, in order to exclude any between-group baseline differences in gene expression. The Weighted Gene Co-expression Network Analysis (WGCNA) package in R (17, 18) was used to identify modules of co-expressed genes. Gene functional enrichment analyses were performed using WebGestaltR (50).

## Data Availability

Requests for data sharing should be made directly to the Principal Investigator, Dr. Don Sin

## ACKNOWLEDGEMENTS

The authors acknowledge the volunteers who participated in the DISARM trial, and Mr. Ryan vander Werff at UBC Biomedical Research Centre for conducting and advising on RNA sequencing. The authors would like to thank Mr. Irving Ding and Mrs. Chun Hong Tao for their financial support of the St Paul’s Foundation COVID-19 Response Fund. SFvE is the Canadian Institute for Health Research/Glaxo Smith Kline (CIHR/GSK) Professor in Chronic Obstructive Pulmonary Disease. DDS holds the De Lazzari Family Chair at HLI and is a Tier 1 Canada Research Chair in COPD.

## AUTHOR CONTRIBUTIONS

### Data acquisition

SM, FSSLF, TS, SFVE, JML, SL, DDS

### Data analysis and interpretation

SM, XL, CXY, AIHC, FSSLF, CWTY, JML, DDS

### Drafting of manuscript

SM, XL

### Critical revision of draft manuscript

SM, XL, CXY, AIHC, FSSLF, CWTY, SFVE, JML, SL, DDS

All authors reviewed and approved the final version of the manuscript.

All authors agree to be accountable for all aspects of the work including data integrity.

## ONLINE SUPPLEMENTARY MATERIAL

This article has an online data supplement.

## DATA SHARING STATEMENT

Requests for data sharing should be made directly to the Principal Investigator, Dr. Don Sin.

## LEGENDS FOR FIGURES APPEARING IN ONLINE SUPPLEMENT

**Figure E1:**
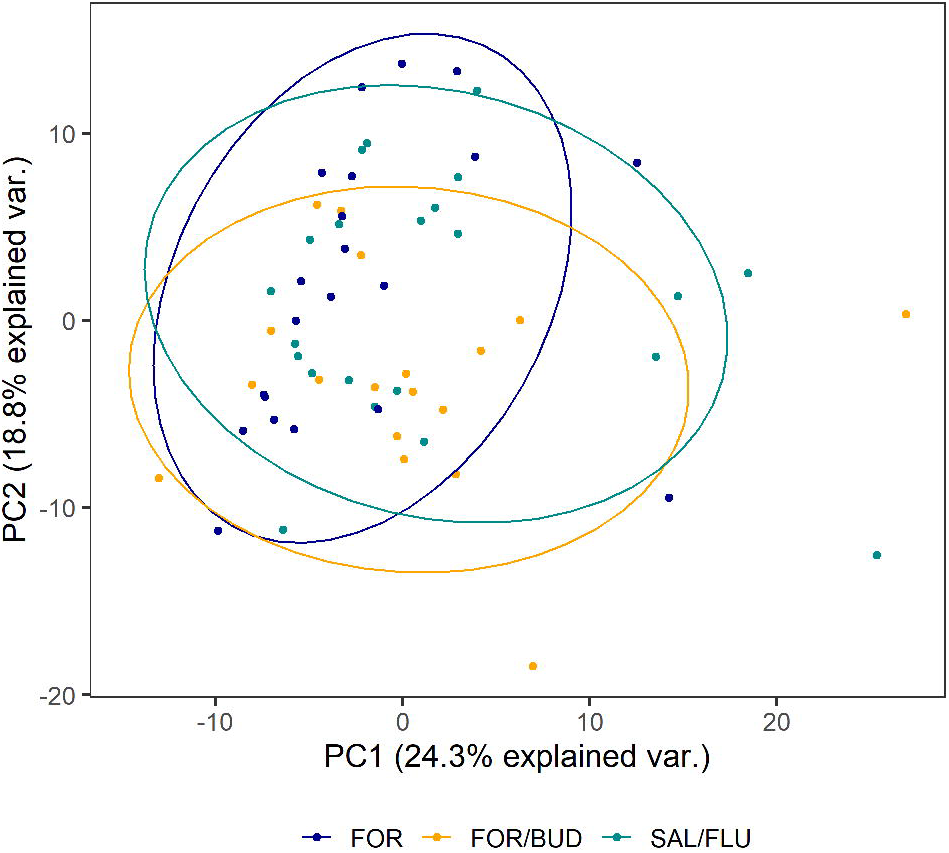
Principal component analysis (PCA) of baseline whole-transcriptome gene expression data. PCA plot grouped by treatment group (n = 61 participants with pre-treatment sequencing data available) shows no obvious separation, suggesting that pre-treatment gene expression was similar between groups. Only genes passing quality control filtering were analyzed.

**Figure E2:**
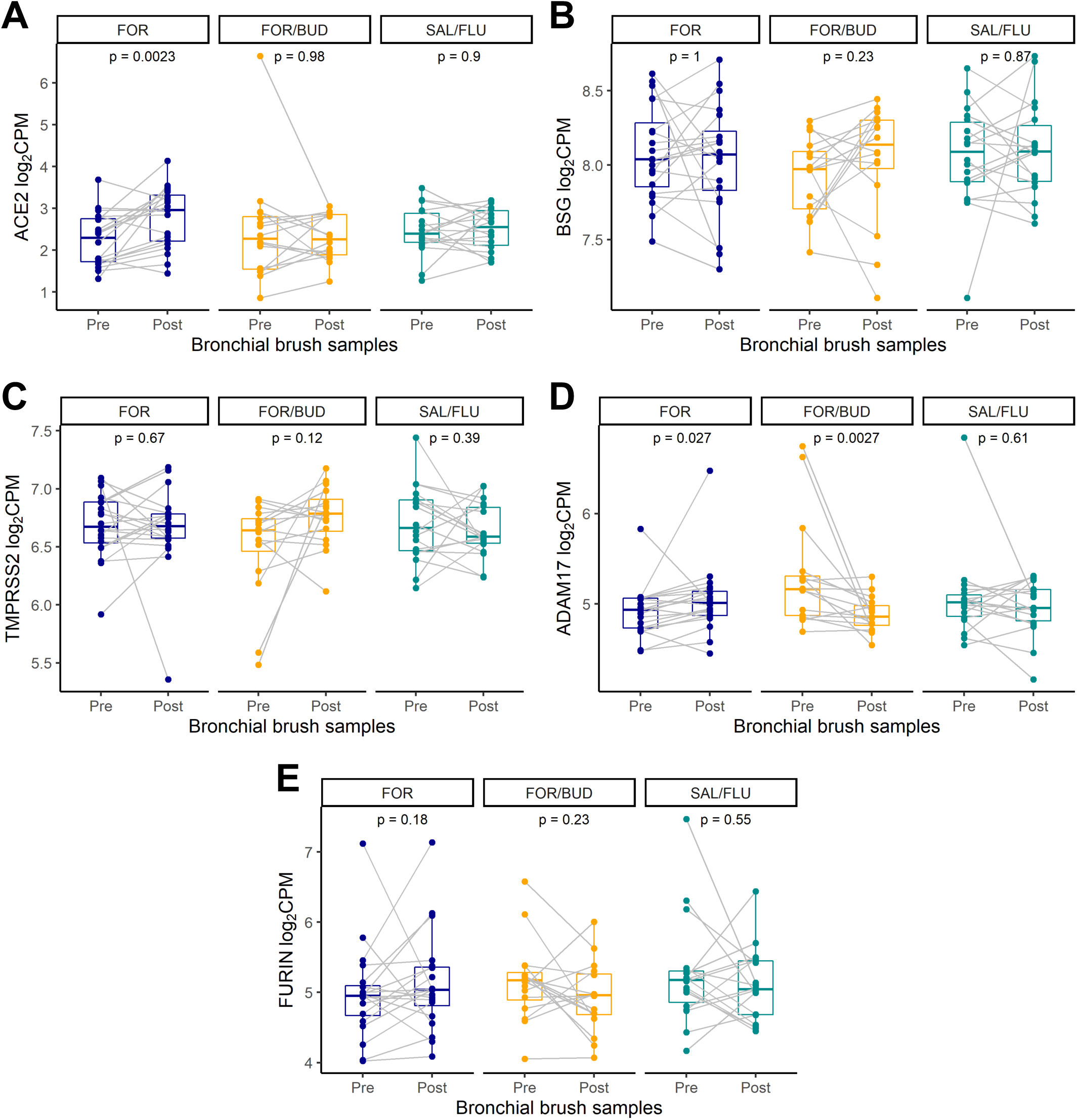
Pre- to post-treatment change in expression (Δlog_2_CPM) of key SARS-CoV-2-related genes. (A) *ACE2*, (B) *BSG*, (C) *TMPRSS2*, (D) *ADAM17*, (E) *FURIN*. Boxplots show median and interquartile range. P value from Wilcoxon signed rank test. Abbreviations: CPM, counts per million.

**Figure E3:**
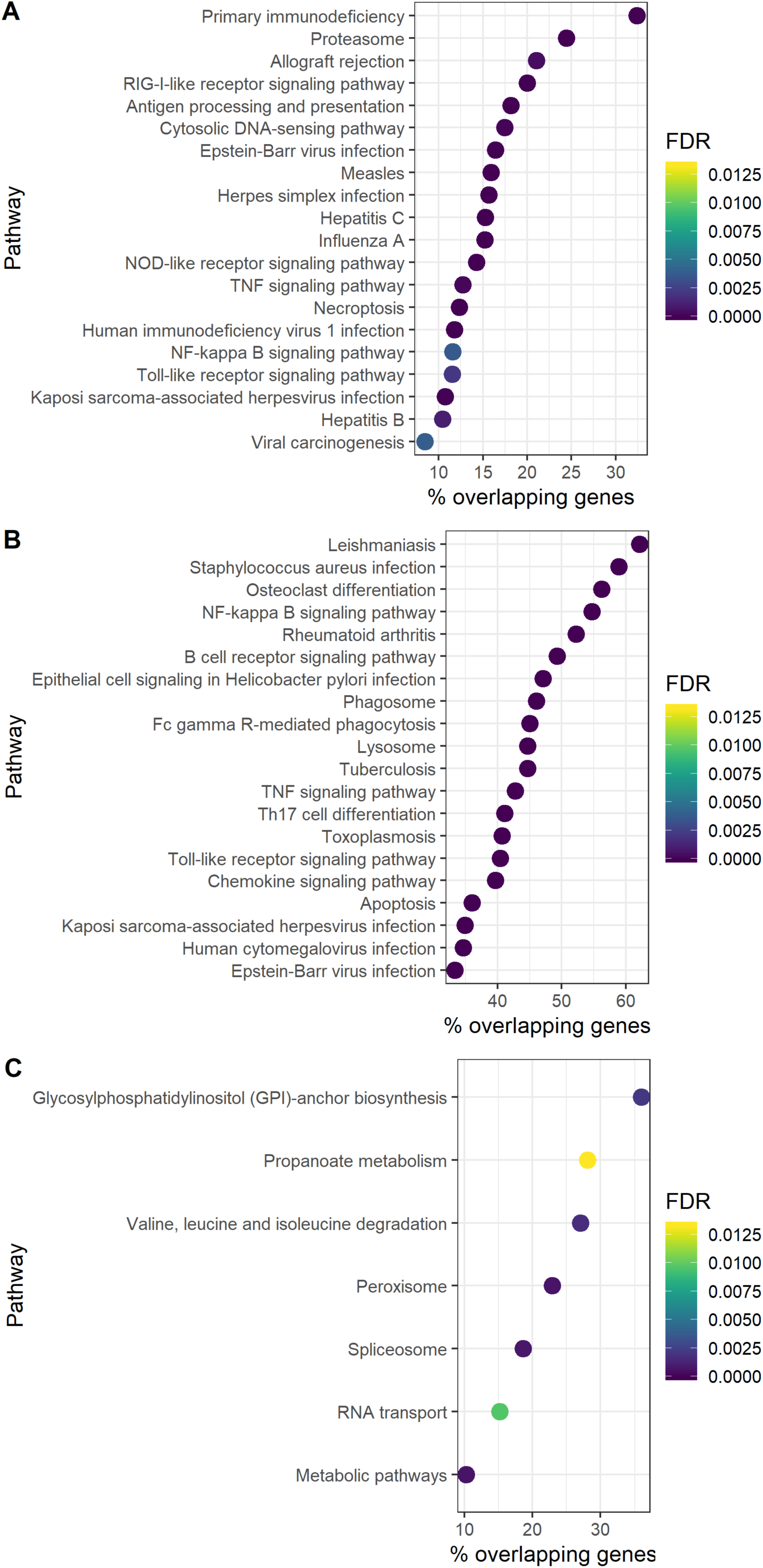
Kyoto Encyclopedia of Genes and Genomes (KEGG) pathway enrichment in SARS-CoV-2-related gene co-expression modules. Top 20 KEGG pathways by FDR are shown, ordered by % overlapping genes (proportion of genes in the KEGG pathway that also appear in the gene module). The *ACE2* module (A) was enriched with genes related to viral diseases including influenza A and measles, as well as disorders of the immune system including primary immunodeficiency and allograft rejection. The *ADAM17*/*FURIN* module (B) was enriched with genes related to bacterial diseases including leishmaniasis and Staphylococcus aureus infection, as well as the autoimmune disease rheumatoid arthritis. The *BSG*/*TMPRSS2* module(C) was not enriched for any human disease pathways at FDR< 0.05. Abbreviations: FDR, false discovery rate.

**Figure E4:**
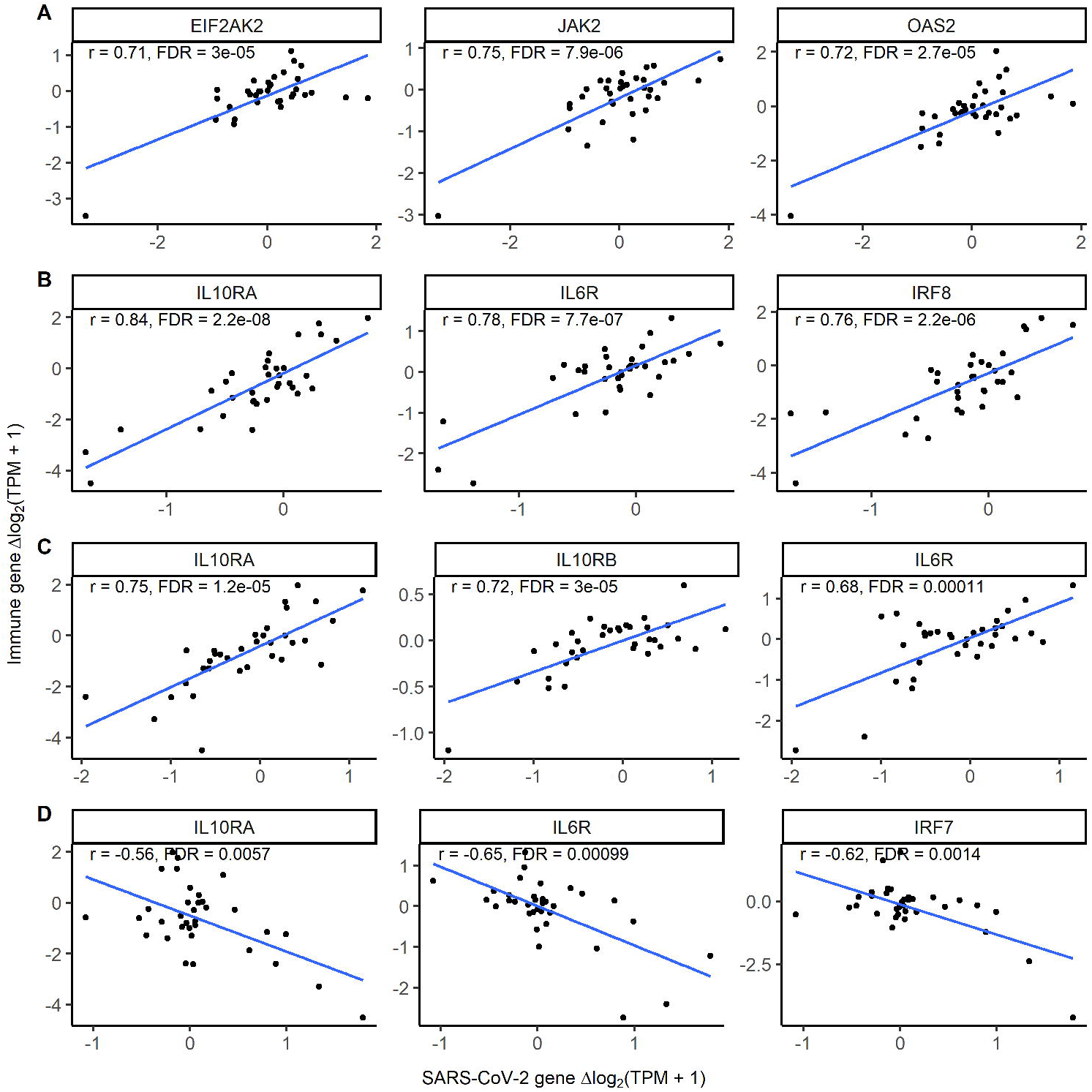
Scatter plots showing correlation of pre- to post-treatment change in selected immune response genes with change in key SARS-CoV-2-related genes for LABA/ICS treatment groups combined. Pre- to post-treatment change quantified as Δlog_2_(TPM+1). r from Pearson correlation. Top 3 correlated innate immune response genes for each SARS-CoV-2-related gene shown: (A) *ACE2*, (B) *ADAM17*, (C) *FURIN*, (D) *TMPRSS2*. There were no correlations with change in *BSG* at FDR< 0.05.

